# Advancing Personalized Cancer Therapy: *Onko_DrugCombScreen* - A Novel Shiny App for Precision Drug Combination Screening

**DOI:** 10.1101/2024.06.20.24309094

**Authors:** Jingyu Yang, Meng Wang, Jürgen Dönitz, Björn Chapuy, Tim Beißbarth

## Abstract

Identifying and validating genotype-guided drug combinations for a specific molecular subtype in cancer therapy represents an unmet medical need and is important in enhancing efficacy and reducing toxicity. However, the exponential increase in combinatorial possibilities constrains the ability to identify and validate effective drug combinations. In this context, we have developed *Onko DrugCombScreen*, an innovative tool aiming at advancing precision medicine based on identifying significant drug combination candidates in a target cancer cohort compared to a comparison cohort. *Onko DrugCombScreen*, inspired by the Molecular Tumor Board (MTB) process, synergizes drug knowledge-base analysis with various statistical methodologies and data visualization techniques to pinpoint drug combination candidates. Validated through a TCGA-BRCA case study, *Onko DrugCombScreen* has demonstrated its proficiency in discerning established drug combinations in a specific cancer type and in revealing potential novel drug combinations. By enhancing the capability of drug combination discovery through drug knowledge bases, *Onko DrugCombScreen* represents a significant advancement in personalized cancer treatment by identifying promising drug combinations, setting the stage for the development of more precise and potent combination treatments in cancer care. The *Onko DrugCombScreen* shiny app is available at https://rshiny.gwdg.de/apps/onko_drugcombscreen/. The Git repository can be accessed at https://gitlab.gwdg.de/MedBioinf/mtb/onko_drugcombscreen.

## Introduction

Cancer treatment is an intricate field, with the ongoing quest to develop therapies that effectively target the disease while minimizing side effects. The application of synergistic drug combinations builds on the idea of lowering the concentration of both drugs to achieve the same effect with fewer side effects and represents an important advancement in cancer therapy [9]. This approach aims to overcome the limitations of single-agent treatments by improving efficacy and reducing the likelihood of drug resistance [7]. However, the task of identifying optimal drug combinations is complicated by the significant variability in tumor types and patient responses, along with the complexities of cancer biology, the high dimensionality of data, and the number of drug combinations far beyond what is possible for clinical testing [18, 23]. These challenges make it difficult to predict which combinations are most likely to be effective in specific cancer molecular type cohorts compared to other molecular subtypes, necessitating advanced computational tools and extensive experimental validation to navigate the vast landscape of potential drug combinations and tailor treatments to cohort patients’ needs [27].

Molecular Tumor Boards (MTBs) are crucial in personalizing cancer treatment, integrating multidisciplinary expertise to interpret genetic data and guide treatment decisions based on a patient’s unique tumor characteristics [25, 21, 19]. Drawing inspiration from the methodologies employed by MTBs, the utilization of drug databases alongside detailed drug’s level of evidence information emerges as a crucial strategy in advancing patient-specific treatment recommendations [25, 21, 19]. This approach not only facilitates the identification of the most suitable drugs for individual patients based on their genetic profiles but also sets the foundation for drug combination prediction based on patient cohorts. Applying statistical methods based on drug databases to the set of recommended drugs for these patient cohorts enables researchers and clinicians to predict more accurately effective drug combinations. This strategy underscores a significant shift towards a data-driven and evidence-based framework to optimize combination therapy for cancer patients, leveraging the increasingly available genetic and pharmacological data to enhance treatment efficacy and patient outcomes.

In recent decades, an increasing application of computational approaches has been developed for the prediction of drug combinations and their effects. Preuer et al. developed *DeepSynergy*, a deep learning-based approach that accurately predicts drug combination synergies for cancer treatments, significantly surpassing traditional performance methods [26]. Similarly, Wang et al. introduced *DeepDDS*, a deep learning model that employs graph neural networks and attention mechanisms to precisely predict and prioritize synergistic drug combinations for cancer treatments, achieving the advantage of enhanced interpretability through chemical substructure analysis [33]. Cheng et al. demonstrated that a network-based methodology, concentrating on the relative configuration of drug–target modules in connection to disease modules, can effectively prioritize potentially efficacious drug combinations for complex diseases such as cancer [5]. *GAECDS*, presented by Li et al., is an innovative approach combining graph autoencoders and convolutional neural networks to accurately predict drug synergy, showing superior performance in identifying efficacious drug combinations [20]. Concurrently, numerous classical machine learning models have also exhibited performance comparable to deep learning methods, demonstrating their robustness and utility in this complex domain. Gayvert et al. showcased that a Random Forest model, utilizing single drug dose responses as features, could accurately predict drug pair synergy and effectiveness in mutant *BRAF* melanomas [11]. Janizek et al. introduced *TreeCombo*, an XGBoost-based approach that leverages the power of gradient boosting to improve predictive accuracy, outperforming DeepSynergy by using drug physiochemical features and cancer cell line gene expression data. The use of XGBoost, which combines multiple decision trees to make robust predictions, demonstrated comparable efficacy to deep learning on medium-scale datasets, while offering the additional benefits of reduced complexity in hyperparameter tuning and enhanced interpretability through *TreeSHAP*, a feature attribution method that identifies the contribution of each variable in a clear and consistent manner [17]. However, current pre-clinical screenings primarily focus on the synergistic effects of drug combinations, often overlooking key factors for clinical success such as potential toxicity and selective efficacy against tumors [18]. At the same time, there is a clear lack of innovative computational solutions to demonstrate their feasibility and benefits in translational applications, especially in the field of cancer, where there is an urgent need to identify combination therapies suitable for specific cancer group patients based on patient-specific biomarkers [6, 29].

In this paper, we present an *Onko DrugCombScreen* Shinyapp designed to address this gap in cancer therapy which could predict the significant drug combination candidates based on the target patient cohort statistical analysis against the comparison cohort. The primary goal of *Onko DrugCombScreen* extends beyond merely providing treatment recommendations based on drug databases like GDKD [8], CiVIC [13], and OncoKB [4]. It integrates statistical methods and data visualization to analyze target cancer type cohort and comparison cohort genetic data against extensive drug databases, thereby uncovering potential drug combinations and mapping them onto cell line data, providing a robust basis for clinical drug screening. Based on the drug evidence levels in the knowledge database for medications, one can directly ascertain whether the variant mapping drugs are selective at the cancer types in the target patient cohort, and the previous studies collected in the database can save workload on drug toxicity analysis. This brings renewed hope for the clinical translation of cancer-type-specific drug combination therapies.

**Table 1.**
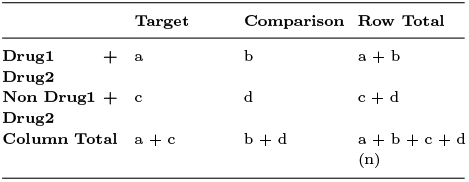
Contingency table for Fisher exact test analysis. In this table, *a* represents the number of patients in the target tumor cohort receiving Drug1+Drug2 co-recommendation, *b* is the number of comparison cohort patients receiving Drug1+Drug2 co-recommendation, *c* is the number of patients in the target tumor cohort not receiving Drug1+Drug2 co-recommendation, and *d* is the number of comparison cohort patients not receiving Drug1+Drug2 co-recommendation.

## Methods

### Fisher’s exact test in subtype recommendations

Besides predicting single drugs, clinicians and researchers are interested in determining whether two drugs are simultaneously recommended for the target tumor type and exhibit significant differences compared to the comparison tumor group. Here, we defined co-recommended drugs as candidate drug combinations that are presented in the `Drug_comb` column in the DrugComb analysis table. We then counted the number of patients in the target tumor cohort in the comparison cohort for each candidate drug combination. Subsequently, we used these four counts to construct a contingency table (Table **??**) and performed a Fisher’s exact test for each candidate drug combination. By analyzing the p-value and odds ratio that are circled with a red rectangle in Figure 1 results obtained from Fisher’s exact test, we can determine if the occurrence of a candidate drug combination is significantly different and assess the magnitude of this difference. The p-values were adjusted using the Benjamini-Hochberg method, as reflected in the `adjust_p.value` column, to account for multiple hypothesis testing and control the false discovery rate (FDR). Additionally, we report drug combination candidate recommendations for cell line data in the final four columns in the DrugComb analysis table to assist with wet-lab validation. (Fig. 1).

**Fig. 1.**
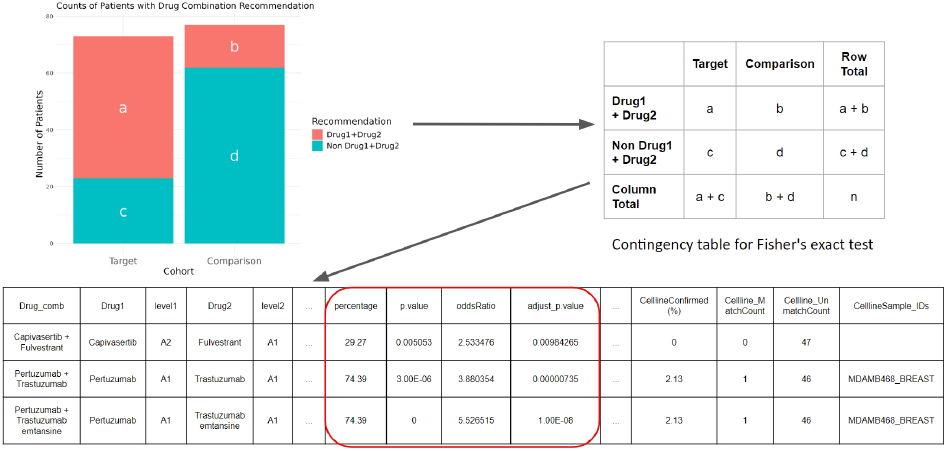
A ‘drug co-recommendation’ refers to the recommendation of two drugs for a single patient, exemplified as Drug1 + Drug2. The bar plot illustrates the counts of such co-recommendations within the target cancer subtype cohort compared to the comparison cancer subtype cohort, leading to the construction of the contingency table on the right. Fisher’s exact test was applied to each drug co-recommendation to assess statistical significance, resulting in the DrugComb analysis table displayed below. The percentage column (`percentage`) indicates the proportion of drug co-recommendations, with the p-value and adjusted p-value columns (`adjust_p.value`, `adjust_p.value`) reflecting the significance level. The odds ratio (`oddsRatio`) provides a measure of the effect size in comparison to the control group. The final four columns offer details on the cell line drug recommendation status, including specific cell line IDs (`individual_id`) used for wet lab validation.

Here, Fisher’s exact test serves as a robust statistical method to determine the significance of the association between the candidate drug combination and the tumor type. Fisher’s exact test is particularly suitable for small sample sizes and for datasets where the assumptions of chi-squared tests are not met.

The null hypothesis for Fisher’s exact test posits that there is no association between the drug combination and the tumor type. Under this assumption, the test calculates the p-value, which represents the probability of observing the current distribution of patients, or a more extreme distribution, if the null hypothesis were true. The p-value is computed using the hypergeometric distribution formula:

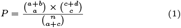

Here, *a, b, c*, and *d* are the cell counts in the 2 *×* 2 table, and *n* is the total sample size (*a* + *b* + *c* + *d*). The *p*-value is then the sum of probabilities (*P*) for all arrangements of the table where the association between the rows and columns is as extreme as or more extreme than in the observed table. A low *p*-value indicates that the observed distribution is unlikely under the null hypothesis, suggesting a significant association between candidate drug combinations and the tumor type.

The odds ratio is calculated as 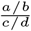, representing the odds of the target tumor group receiving Drug1+Drug2 compared to the odds of the comparison group receiving the same. An odds ratio greater than 1 indicates a higher likelihood of the target cohort receiving the drug combination compared to the comparison cohort, while an odds ratio less than 1 suggests the opposite.

### Drug Level of Evidence

Here, we adopted the MTB drug’s level of evidence category approach proposed by Perera-Bel [25]. As shown in Table 2. “*A*” signifies evidence for the same cancer type, while “*B* “ indicates evidence for any other cancer type. Horizontally, Level 1 represents evidence supported by regulatory agencies or clinical guidelines. Level 2 includes evidence from clinical trials. Finally, Level 3 consists of preclinical trial evidence. Therefore, based on the different target cancer types of drugs and their respective clinical evidence, six levels of drug evidence are derived: A1, A2, A3, B1, B2, B3. With this drug level of evidence, the selection of recommended drugs for specific cancer types and their clinical strength can be clearly defined, which can guide the clinical decision.

**Table 2.**
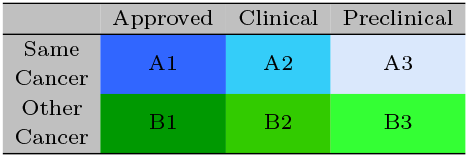
Drugs obtained from the Drug knowledge database are classified into clinically relevant categories using a system of six levels of evidence.

|  | Approved | Clinical | Preclinical |
| --- | --- | --- | --- |
| Same Cancer | A1 | A2 | A3 |
| Other Cancer | B1 | B2 | B3 |

### Material

#### Analysis tools

*Onko DrugCombScreen* was implemented using R (v.4.3.1) and R Shiny (v.1.8.0). This shiny app integrated a variety of R programming language packages for comprehensive bioinformatics analysis. For parsing and generating data structures, we utilized readxl v1.4.1 [38, 37]. To facilitate data manipulation and transformation, we employed packages like reshape2 v1.4.4 [36], tidyr v1.2.1 [37], and dplyr v1.0.10 [39]. We applied packages such as maftools v2.12.0 [22], clusterProfiler v4.4.4 [41], and VariantAnnotation v1.42.1 [24] for the analysis of somatic variants, functional profiles of genes. For data visualization, we used packages such as circlize v0.4.15 [15] for circular visualizations, ggalluvial v0.12.3 [3, 2] for alluvial diagrams, ggrepel v0.9.2 [28] for label clarity, ComplexHeatmap v2.12.1 [14] for sophisticated heatmaps, and ggplot2 v3.4.0 [31] for creating customizable static plots.

#### Data Source

The harmonized drug database, is derived from open-source drug knowledge databases including GDKD [8], CiVIC [13], and OncoKB [4], utilizes the *DrugBank Vocabulary* dataset from DrugBank [40] to standardize drug synonyms. TCGA-BRCA data and breast cancer cell line data used in the case study were collected from UCSC Xena hubs [12].

## Results

### Case Study: Application and Validation Using TCGA-BRCA Data

#### Dataset Selection and Processing

In this case study, the *Onko DrugCombScreen* was applied to the TCGA-BRCA dataset to validate its efficacy in identifying effective drug combinations for breast cancer. The TCGA-BRCA dataset, derived from the TCGA Pan-Cancer (PANCAN) initiative, was chosen for its comprehensive genetic profiling, including extensive data on copy number variations (CNVs), single nucleotide variations (SNVs), and molecular subtype profiles [35]. This dataset provides a broad coverage of genetic variations, making it an ideal resource for this analysis. Additionally, cell line data from the Cancer Cell Line Encyclopedia (Breast) were incorporated to complement the analysis [1]. All of the above data set are available in UCSC Xena hubs [12].

In the preprocessing phase, somatic mutation data from the TCGA Pan-Cancer (PANCAN) was converted into a compatible CSV file for analysis by the *Onko DrugCombScreen*. This process involved filtering the dataset to isolate BRCA cancer data and further stratifying it into molecular subtypes - Luminal A/B, HER2+, Normal-like, and Basal-like. In this case study, we used Normal-like breast cancer as the comparison cohort, and HER2+, and Basal-like subtypes as the target cohorts, respectively, to analyze and validate the efficacy of *Onko DrugCombScreen*. Additionally, by integrating cell line data, *Onko DrugCombScreen* provided guidance on suitable cell lines for subsequent experimental validation.

#### Validation And Results

The drug co-recommendation comparison analysis revealed significant disparities between the three BRCA subtypes (HER2+ and Basal-like) and the Normal-like BRCA data. Significant drug co-recommendations extracted from *Onko DrugCombScreen* were compared with combinational therapies in Wang’s review [34], as well as FDA-approved drug combinations to validate the effectiveness. As the supplementary table A shows, the “adjust p.value” and “OR” (odds ratio), obtained from the Fisher’s exact test, indicate the significance and magnitude of the drug combination in the target cohort compared to the comparison cohort, the “Percentage” depicts the proportion of the drug combination recommended in the target cohort. Setting the threshold at *adjust p*.*value* < 0.05, *OR* > 1, and *Percentage* > 50% retains around 30% of the significant candidate drug combinations (30250/111987 in HER2+ vs Normal-like and 48348/112069 in Basal-like vs Normal-like). Notably, these stringent criteria preserved almost all approved and clinical trial drug combinations, including the approved combinational therapy of Pertuzumab + Trastuzumab for the HER2+ subtype and Pembrolizumab + Paclitaxel for Triple-Negative Breast Cancer (TNBC). These results highlight *Onko DrugCombScreen*’s accuracy in identifying clinically relevant drug combinations, confirming its effectiveness. Besides, upon comparison with the DrugComb.org database [42], it was found that none of the approved and currently in clinical trial drug combinations of breast cancer had any recorded synergy scores.

The validation analysis demonstrates that the *Onko DrugCombScreen* is adept at identifying established breast cancer drug combinations in the BRCA subtypes like HER2+ and Basal-like when compared to Normal-like BRCA subtype. This finding not only validates the tool’s effectiveness but also highlights its potential in discovering novel drug combinations for various cancer types. Consequently, the case study accentuates the utility of the *Onko DrugCombScreen* in providing targeted and efficacious drug recommendations.

### Data Analysis Workflow of Onko DrugCombScreen

The data analysis workflow of *Onko DrugCombScreen* is depicted in Figure 2: Variant data such as single nucleotide variants (SNV) and copy number variants (CNV) from both the cancer subtype cohort and the comparison cancer subtype cohort are preprocessed and converted into variant tables compatible with *Onko DrugCombScreen*. These patient variant data are then mapped to public drug databases (CiVIC, GDKD, OncoKb) after integration with variant interpretation annotations and drug evidence levels for drug recommendations. The resulting drug recommendations are subjected to statistical analysis, focusing on the statistical differences in drug combination candidates observed between the target cancer subtype group and the comparison cancer subtype group. To identify drug combination candidates that are significantly and frequently recommended in the target group compared to the comparison group, Fisher’s exact test is applied. Subsequently, the selected drug combination candidates undergo an integrated analysis with cell line data to identify available cell line samples, facilitating wet-lab validation. Additionally, all analysis results are visualized, making the findings clearer and more intuitive. The integration of these processes is crucial for confirming drug combination recommendations for the cancer type of interest. The final validation stage may include conducting wet-lab drug screenings to confirm the analysis results and deepen the understanding of the underlying biological mechanisms (Fig. 2).

**Fig. 2.**
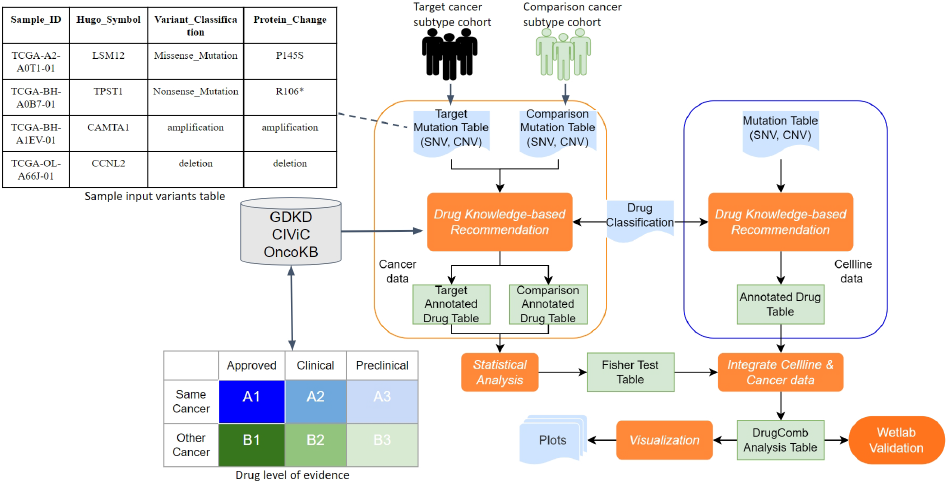
The workflow for *Onko DrugCombScreen* drug combination data analysis. After the recommendation process based on drug knowledge, SNVs and CNVs are merged into an annotated drug table. Following statistical analysis and integration of cell line data, the final DrugComb analysis table will be used for visualization and wet lab validation.

#### Data Preprocessing

SNVs and CNVs are typically stored in formats such as VCF, MAF, TXT, or Excel. A preprocessing step is necessary to convert these various formats into csv format (Fig 2). These dataframes are then suitable for use in the knowledge-based drug recommendation analysis within *Onko DrugCombScreen*.

#### Matching Rule Between Variant Annotations in Patients’ data and Database

Due to the different annotation descriptions of variants in the three-drug databases (GDKD [8], CiVIC [13], and OncoKB [4]) and original patients’ variants data, we harmonized the three drug databases and designed a matching rule based on the interpretation of biological significance (Table 3). All variant classes or effects map to the biological interpretations of “loss”, “gain”, or “mutation”. We can then associate the original variants (Table 4) with the information in the knowledge database based on biological interpretations and obtain the relevant target drug information.

**Table 3.** Biological interpretation of variant annotations in drug databases.

| DB | variant | Interpretation |
| --- | --- | --- |
| GDKD/CIVIC/OncoKB | “splice” | loss |
| GDKD/CIVIC/OncoKB | “delins” | loss |
| GDKD/CIVIC/OncoKB | “ins” | ins |
| GDKD/CIVIC | “del” | del |
| GDKD | “indel” | loss |
| GDKD/CIVIC | “fs” | loss |
| GDKD/CIVIC/OncoKB | “deletion” | loss |
| GDKD/CIVIC/OncoKB | “amplification” | gain |
| GDKD | mut | mutation |
| GDKD | any | mutation |
| CIVIC | loss/loss-of-function | loss |
| CIVIC | “mutation” | mutation |
| CIVIC | “^expression” | gain |
| CIVIC | “Overexpression” | gain |
| CIVIC | “Underexpression” | loss |
| OncoKB | Truncating Mutations | loss |
| OncoKB | Oncogenic Mutations | mutation |
| CIVIC | “FRAMESHIFT” | loss |
| CIVIC | “FRAME SHIFT” | loss |
| OncoKB/CIVIC | Exon 17 mutations | mutation (exact match) |
| CIVIC | Exon 19 Deletion | loss (exact match) |
| CIVIC | EXON 14 SKIPPING MUTATION | mutation (exact match) |

**Table 4.**
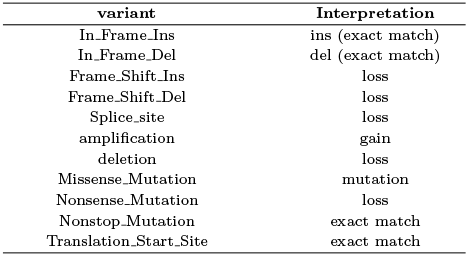
Biological interpretation of variant annotations in patients’ variants.

| variant | Interpretation |
| --- | --- |
| In_Frame_Ins | ins (exact match) |
| In_Frame_Del | del (exact match) |
| Frame_Shift_Ins | loss |
| Frame_Shift_Del | loss |
| Splice_site | loss |
| amplification | gain |
| deletion | loss |
| Missense_Mutation | mutation |
| Nonsense_Mutation | loss |
| Nonstop_Mutation | exact match |
| Translation_Start_Site | exact match |

#### Drug Recommendation Annotation

After the matching rule is defined, the drug knowledge-based analysis was performed to export the drug recommendation tables across all target and comparison subtype data. However, due to discrepancies in drug nomenclature across the three drug databases, we employed the “DrugBank Vocabulary” dataset from DrugBank to standardize synonymous drug names. Subsequently, each drug name was annotated to its final drug class. This annotation is stored in the columns “Origin Drug Name” and “Classified Drug Name” of the DrugComb analysis table. Additionally, other useful information such as variant type is annotated in the “mutType” column, and variant match status—which indicates whether the amino acid change in the raw data exactly matches the database records or not—is saved in the “Match Sign” column.

#### Data visualization

*Onko DrugCombScreen* provides a variety of charts for visual analysis results, allowing users to understand data more intuitively (Fig. 3). The application integrates multiple plotting functions, including volcano plots, heatmaps, circle plots, alluvial diagrams, upset plots, and bar charts. These visualization tools help to easily identify recommended drugs or candidate drug combinations for subsequent wet-lab analysis and validation. Users can configure settings in the left panel of *Onko DrugCombScreen* and customize the resolution for PDF file export.

**Fig. 3.**
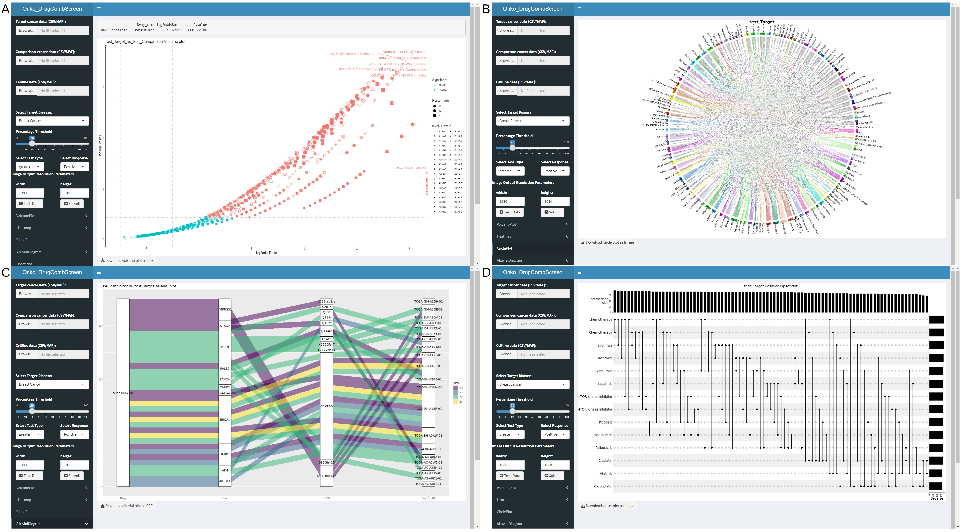
Visualization of the *Onko DrugCombScreen*. (A) Volcano plot identifying significant drug combinations. (B) Circle plot depicting the most proportional drug co-recommendations. (C) Alluvial diagram tracing mutations back to recommended single drugs. (D) UpSet plot showing the top-recommended single drugs and their intersections.

## Discussion

Drug combinations are widely recognized for their benefits in cancer therapy. Here, we developed the *Onko DrugCombScreen* shiny app integrated statistical analysis to identify the most significant candidate drug combinations for a target tumor type cohort. We utilized drug knowledge base recommendations derived from mutation data of the targeted cancer patient cohort and the comparison cohort to identify drug co-recommendations. This is complemented by integrating cell line data to assist in the validation of biological experiments. *Onko DrugCombScreen*’s ability to identify effective drug combinations, as demonstrated in the TCGA-BRCA case study, suggests a promising way toward more tumor-type tailored and effective cancer combination therapy.

In contrast to current computational methods that mainly focus on synergy and dose-response matrices, *Onko DrugCombScreen* is a drug knowledge-based analysis approach. It not only provides therapeutic recommendations but also offers guidance for clinical research, thereby integrating more closely with clinical applications. Moreover, all drug recommendations can be traced back to patient genetics and variants through the visual alluvial diagram of *Onko DrugCombScreen*. Utilizing the drug database which is also employed by the MTB report, each recommended drug’s level of evidence and response is explicitly defined. This clarity effectively aids in addressing issues of selectivity in the recommended drug combinations, issues that are often overlooked in previous methods. Moreover, the utilization of drug databases for the recommendation of candidate drug combinations based on patient gene mutation profiles can potentially reduce the effort required for toxicity analysis [32, 10]. These databases provide valuable information on the relationships between individual drugs and specific gene mutations or molecular targets, which can guide the selection of drug combinations with potentially favorable efficacy profiles. Furthermore, the drugs included in these databases are often approved or under clinical trials, meaning that their toxicity profiles have been extensively studied and characterized [16, 30]. This existing safety data can serve as a foundation for assessing the toxicity of drug combinations, as it provides insights into the common adverse events, dose-limiting toxicities, and recommended dosing schedules of the individual drugs. By leveraging this information, researchers can streamline the toxicity assessment process and make more informed decisions when designing drug combination studies. However, it is crucial to acknowledge that the toxicity of a drug combination may not be a simple sum of the individual drug toxicities and thorough safety assessments of the specific combination, considering factors such as drug-drug interactions, dosing, scheduling, and special patient populations, are still necessary to ensure a comprehensive understanding of the toxicity profile.

Looking forward, the potential for further development of the *Onko DrugCombScreen* is substantial. Due to the lack of data on synergy and dose specificity in the drug database, *Onko DrugCombScreen* is currently unable to provide such information. In future iterations, it is feasible to incorporate synergy and dose specificity data. Moreover, advancements in artificial intelligence and machine learning can be leveraged to enhance data analysis based on drug knowledge, thereby continuously improving the predictive accuracy and relevance of the tool.

## Conclusion

In conclusion, the *Onko DrugCombScreen* Shiny app represents an innovative tool in the field of precision cancer therapy, offering a novel drug knowledge-based approach to drug combination screening. Harnessing the power of drug knowledge database analysis, this application integrates advanced statistical analysis and data visualization techniques to explore and identify effective drug combinations. It effectively utilizes drug recommendations from targeted cancer cohort and comparison cohort, combined with cell line data, to provide prominent drug co-recommendations for targeted cancer type. Validated through a TCGA-BRCA case study, the application has demonstrated its potential in accurately identifying both existing and novel drug combinations, aligning with the evolving field of precision oncology. Looking ahead, the integration of artificial intelligence and machine learning technologies holds the promise of further enhancing its predictive capabilities, making it a valuable tool in the quest for more targeted and effective cancer treatment approaches.

## Data Availability

All data produced are available online at TCGA repository

## Supporting information

**Supplementary File S1: Sample data files for Onko DrugCombScreen testing**

This file includes three CSV files:

- `Primary_Her2_mutationSNVCNV DF.csv`: Target cancer subtype data.
- `Comparison_Normallike_BRCA_mutation DF.csv`: comparison cancer subtype data.
- `Cellline_CCLEBRCA_Mutation_DF.csv`: Cell line data.

**Supplementary Table S1. Comparison of Current Approved Therapies and Clinical Trial Combinations for Breast Cancer Treatment with Onko DrugCombScreen Based Screening result [34]**.

## Author contributions statement

JY, MW, BC, JD, and TB designed the study. JY and BC provided major contributions to the computational method and visualization. JY designed, developed, and implemented the Shiny app. MW tested the Shiny app and helped identify problems and bugs. JY wrote the manuscript. All authors critically reviewed the content and approved the final manuscript.

## Acknowledgments

The authors would like to acknowledge the Volkswagen Foundation’s support in MTB-report project. JY was supported by the Ph.D. program “Genome Science” - International Max Plank Research School, part of the Göttingen Graduate Center for Neurosciences, Biophysics, and Molecular Biosciences. JD and TB are members of the Göttingen Campus Institute Data Science.

